# Tracking Neural, Sensory, and Sensorimotor Adaptation to Progressive Vision Loss in Inherited Retinal Dystrophies: A Multimodal Longitudinal Study Protocol

**DOI:** 10.64898/2026.08.18.26360630

**Authors:** Alessia Verroca, Elena Franchin, Sonia Mele, Ilaria Siviero, Isolde Martina Busch, Anna Benamati, Javier Sanchez Lopez, Chiara Quisisana, Angelica Filosa, Valerio Marino, Leonardo Colombo, Paola Cesari, Michela Rimondini, Daniele Dell’Orco, Maria Paola Cecchini, Chiara Mazzi, Silvia Savazzi

## Abstract

Individuals with inherited retinal dystrophies (IRDs) undergo a slow, genetically heterogeneous loss of vision, yet how the visual cortex and non-visual sensory, motor, and psychological systems adapt to this deprivation remains poorly characterized. Existing evidence comes mainly from single-modality, cross-sectional studies that rarely account for genetic heterogeneity, making it hard to distinguish adaptive change from a direct, non-retinal mutation effect, since several IRD genes are not retina-specific. To address this gap, we designed an observational, longitudinal, multimodal protocol that combines ophthalmological, genetic, and in silico characterization with electrophysiological (steady-state visual evoked potentials and TMS-EEG), chemosensory, sensorimotor, and psycho-personological assessments. Patients aged 18 to 75 years with rod-cone (retinitis pigmentosa, Usher syndrome) or cone and cone-rod dystrophies will be assessed at baseline (T0) and at an 18-month follow-up (T1); sighted controls, matched for age, sex, and handedness, will complete the same battery once. Importantly, pairing genotypic with phenotypic data allows changes in non-visual domains to be interpreted against, rather than independently of, each patient’s molecular background. We expect individuals with IRDs to differ from controls in visual cortical responsiveness and in selected non-visual sensory and sensorimotor measures, with genotype-related differences explored where sample size permits. Given the rarity of IRDs, the design is exploratory and emphasizes effect sizes and individual variability over large-sample inference. The protocol was approved by the Ethics Committee of the University of Verona (CARP 08.R1/2024) and follows the Declaration of Helsinki and the GDPR; findings will be disseminated through peer-reviewed publications and shared with patients and IRD patient associations.

## 1. Introduction

### 1.1 Inherited retinal dystrophies as a model of progressive sensory loss

Inherited retinal dystrophies (IRDs) are a group of rare, genetically heterogeneous ocular disorders characterized by progressive degeneration of photoreceptors and subsequent gradual vision loss (Hartong et al., 2006; Duncan et al., 2018). This genetic heterogeneity translates into phenotypic variability, with differences in age of onset, rate of progression, severity of visual impairment, and pattern of retinal degeneration. Beyond the well-characterized alterations at the ophthalmologic level, including reduced visual acuity, progressive visual field constriction, and retinal remodeling (Hartong et al., 2006), considerably less is known about how the central nervous system and other sensory systems adapt to the progressive reduction of visual input.

IRDs provide a particularly informative model for investigating sensory adaptation because visual deprivation may emerge at different stages of life and progress along distinct spatial trajectories. In many IRDs, visual function is relatively preserved during the first years of life, with symptoms emerging later, although some forms are already associated with severe visual impairment at birth or in early infancy (Bock & Fine, 2014; Kolarik & Moore, 2024). In addition, IRDs encompass markedly different degenerative topographies. Rod-cone dystrophies, including retinitis pigmentosa and Usher syndrome, typically produce an initial peripheral-to-central pattern of visual-field loss. In contrast, cone and cone-rod dystrophies predominantly affect central vision at earlier stages, with peripheral visual function remaining relatively preserved until later in the disease course. These complementary patterns make IRDs a useful natural model for examining how the nervous system responds not only to progressive reduction in visual input, but also to differences in the spatial distribution and developmental timing of visual deprivation.

### 1.2 Genetic and Clinical heterogeneity of IRDs as a source of variability in adaptation

IRDs are linked to mutations in genes that affect the visual process at different stages, most often involving early phototransduction, ciliary trafficking, lipid metabolism, and the visual cycle (Wright et al., Nat Rev Genet, 2010; Duncan et al., Transl Vis Sci Technol, 2018). To date, more than 250 genes have been associated with IRDs (RetNet, https://retnet.org/, accessed July 17, 2026), and while the common outcome is photoreceptor cell death, a clusterization of the genetic features of IRD patients is essential both for grounding assessments in a defined molecular background and for laying the basis for future personalized therapies.

Coupling genetic information with high-resolution structural models of the protein expressed by the mutated genes can lead to useful in silico models that permit the assessment of structural and functional consequences of the specific mutations (Marino et al, Human Mol Gen 2018) and the prediction of functional perturbation at a systems level (Dell’Orco et al. Cell Mol Life Sci, 2014). This is especially useful when the goal is to establish the mechanistic effects of different mutations in the same gene, which is often the case with IRDs.

IRDs are heterogeneous not only in their genetic basis but also in the topography of degeneration, and a protocol meant to serve the whole group must accommodate this range. The most prevalent and best-characterized forms of IRDs are the rod-cone dystrophies, in which degeneration is rod-predominant, and the clinical trajectory is consistently reported. Both retinitic pigmentosa (RP) and Usher Syndrome (USH) are classified as rod-cone dystrophies, a category of inherited retinal degenerations characterized by primary dysfunction and progressive loss of rod photoreceptors, followed by secondary cone degeneration (Hamel, 2006; Hartong et al., 2006). This rod-predominant pattern of photoreceptor loss explains the characteristic clinical progression: RP typically presents with night or scotopic blindness (nyctalopia) as the initial symptom, often emerging in childhood or adolescence, followed by progressive peripheral visual field constriction advancing in a characteristic peripheral-to-central gradient pattern, often described as “tunnel vision” (Hamel, 2006; Hartong et al., 2006).

US presents with the same progressive visual deterioration, with the additional complication of congenital or progressive hearing loss, which normally precedes retinal degeneration (Bonnet & El-Amraoui, 2012; Mathur & Yang, 2015), thereby combining audiovisual sensory deprivation with profound implications for multisensory integration and communication abilities.

A second, opposite pattern is seen in the cone and cone-rod dystrophies, in which degeneration begins in the cone-rich macula of the central retina and is accompanied by disabling symptoms such as reduced visual acuity, impaired color vision, photophobia, and a central scotoma. At the same time, the peripheral field is relatively spared until later stages. Rod involvement with night blindness typically follows in the cone-rod subtype (Gill et al., Br J Ophthalmol, 2019). Together, these forms span the range of degeneration topographies the protocol is built to accommodate, from periphery-first to center-first vision loss.

### 1.3 Neural adaptation to progressive vision loss

Progressive retinal degeneration in IRDs offers a valuable model for investigating how the nervous system adapts to reduced sensory input. Unlike congenital or early-onset blindness, late-onset IRDs involve gradual vision loss that unfolds after the visual cortex has undergone structural and functional specialization (Castaldi et al., 2020). As photoreceptors progressively degenerate, reduced retinal input to the visual cortex may create conditions that trigger large-scale neural reorganization.

The nature and extent of neural adaptation in progressive late-onset blindness, such as that characterizing IRDs, remain considerably less investigated than in early-onset blindness. Moreover, the consequences for visual cortical functions remain actively debated, with evidence pointing to substantial differences between early and late blindness in the degree, mechanism, and functional significance of cortical reorganization (Bock & Fine, 2014). In this context, an open question is whether progressive visual deprivation is associated with changes across different functional domains. The timing of disease onset and the rate of progression appear to shape fundamentally different patterns of cortical reorganization, with late-onset progressive conditions such as IRDs occupying a distinct position within the spectrum of visual deprivation models.

### 1.4 Genotype-related effects as an alternative explanation for cross-domain changes

The plasticity described above is not the only possible explanation for cross-domain changes in IRD patients. Some of what appears to be adaptation may instead be a direct pleiotropic effect of the causative mutation, expressed in a tissue other than the retina. Several IRD-causing genes encode proteins that are not retina-specific: the cyclic nucleotide-gated (CNG) channels central to phototransduction, for instance, are also expressed in olfactory sensory neurons, and mutations in CNGB1 or CNGA2 produce measurable olfactory dysfunction alongside retinal degeneration (Charbel Issa et al., 2018; Karstensen et al., 2015; Sailani et al., 2017; Geada et al., 2023). A comparable mechanism has been described for ciliopathy genes such as CEP290, where loss of ciliary G-protein trafficking in olfactory neurons causes anosmia independently of any visual-to-olfactory compensatory process (McEwen et al., 2007). In such cases, a genotype-driven deficit in a non-visual sensory system could be mistaken for cross-modal compensation if genotype is not accounted for. Moreover, recent transcriptomic analyses show that a mutation in a single photoreceptor-specific gene can alter transcript expression across the whole retina, with effects propagating to the visual cortex (Avesani et al., bioRxiv 2026), indicating that direct genetic effects are not necessarily confined to the cell type in which the mutated gene is canonically expressed. We therefore pair multimodal phenotyping with genotypic and in-silico characterization, which allows cross-domain changes to be interpreted in relation to both genetic background and visual disease characteristics. This is also why the domains are measured together rather than in isolation, as most prior work has done: only within the same patients can a change in one channel be read against the others and the underlying mutation. Critically, the potential influence of genetic factors, which may concurrently affect not only retinal but also chemosensory, motor, or cognitive phenotypes, has been largely overlooked.

### 1.5 Rationale for a longitudinal multimodal approach

Most existing studies focus on a single modality (e.g., VEPs, structural MRI) and/or are cross-sectional. Such designs do not allow the longitudinal trajectory of adaptation to be characterized and provide limited information on the relationships among different functional and sensory domains. Moreover, they rarely account for the substantial genetic heterogeneity underlying IRDs, despite growing evidence that distinct mutations, even within the same gene, may differentially shape both disease progression and multidimensional phenotypic expression. To address these gaps, we designed an observational, longitudinal, multimodal study. Our protocol combines detailed ophthalmological and genetic characterization with electrophysiological measures (electroencephalography -EEG- and transcranial magnetic stimulation combined with EEG -TMS-EEG), sensory testing of olfactory and gustatory function, motor/proprioceptive assessments based on kinematics and postural adjustments, and psychological and personality questionnaires. Patients will undergo the multimodal assessment at baseline (T0) and at the 18-month follow-up (T1), whereas healthy controls will be assessed at a single timepoint. Critically, this multimodal phenotyping is coupled with comprehensive genetic screening and in silico biochemical characterization of each patient’s mutations. By examining genotypic profiles together with multidimensional phenotypic trajectories, we aim to move beyond broad diagnostic categories (e.g., “retinitis pigmentosa” or “Usher syndrome”) toward a more individualized characterization of IRD profiles.

### 1.6 Objectives and Hypotheses of the Protocol

The primary objective of this study is to characterize neural, sensory, sensorimotor, and psychological features in individuals with IRDs. At baseline, we will compare the multimodal profile of individuals with IRDs with that of sighted controls. In patients, longitudinal changes from baseline (T0) to the 18-month follow-up (T1) will be examined to characterize changes over the course of the disease. We expect individuals with IRDs to show differences from sighted controls in visual cortical responsiveness and in selected non-visual sensory (i.e., olfactory and gustatory) and sensorimotor measures. Psychological and personality-related factors (e.g., resilience and mindfulness) will also be examined in relation to how individuals experience and adjust to the condition, as well as to patterns of adaptation and behavioral compensation across functional domains.

The secondary objective is to investigate factors associated with individual differences in adaptation to progressive visual impairment. Within the patient group, neural, sensory, and sensorimotor measures will be examined in relation to the clinical characteristics and progression of visual impairment. Given the genetic heterogeneity of IRDs, genotype-related differences will also be explored where sample size permits.

### 1.7 Significance and Impact of the Protocol

This integrative protocol offers the potential to: (i) establish a multimodal framework for characterizing sensory and neural adaptive changes in progressive visual loss due to IRDs; (ii) inform personalized profiling for IRD patients by linking neural, sensory, and psycho-personological features to specific genotypes. Ultimately, a comprehensive approach integrating these multiple dimensions within a unified framework can guide targeted rehabilitation strategies and ultimately improve the quality of life for individuals living with progressive vision loss.

## 2. Methods and Analysis

### 2.1 Study Design

This is an observational, longitudinal, multimodal study with two timepoints: baseline (T0) and an 18-month follow-up (T1). Patients with IRD will undergo a comprehensive assessment battery at both time points. In contrast, healthy controls will undergo the same assessment protocol only once, serving as a cross-sectional reference group. The design is fully exploratory and non-interventional, focusing exclusively on phenotypic profiling rather than treatment.

### 2.2 Setting and Recruitment

Patient recruitment will be conducted in collaboration with the Ambulatory for Hereditary Retinal Dystrophy Studies at St. Paolo’s Hospital, Milan, Italy, under the supervision of Dr. L. Colombo (Ophthalmology Unit). Genetic diagnosis will be required for IRD patients, with reference to the molecular epidemiology established by Colombo et al. (2021), which characterized 591 Italian probands with retinitis pigmentosa and Usher syndrome. Healthy controls will be recruited from community volunteers and matched to patients with IRDs by age, sex, and handedness.

### 2.3 Sample Size & Power Considerations

Given the exploratory nature of the protocol and the lack of comparable prior multimodal studies in IRDs, a formal power calculation is not feasible. The sample size will therefore be determined by the available recruitment capacity and the protocol’s feasibility. The methodological foundation relies on previous literature that predominantly consists of single-case reports or studies with very limited sample sizes, which constrains generalizability; performing a post-hoc power analysis could ensure the correctness of the sample size. Emphasis will be placed on effect size estimation, individual-profile analyses, and longitudinal within-subject changes rather than hypothesis-driven large-sample inferential statistics. The recruited patient cohort will need to be matched with a healthy control group to enable statistical comparison and assessment of the significance of the observed effects.

### 2.4 Eligibility Criteria

Inclusion criteria for patients:

- Age 18–75 years.
- Genetically confirmed diagnosis of an IRD, spanning rod-cone dystrophies (Retinitis Pigmentosa, RP, or Usher Syndrome, USH) and cone or cone-rod dystrophies. Genotypes will be characterized for every patient and used for stratification; the gene panel and the stratification scheme, spanning genes known to be specifically linked to IRDs, will be defined by the biochemistry/molecular genetics team.
- Ability to provide written, informed consent.

Exclusion criteria for patients:

- Presence of major neurological, psychiatric, or metabolic disorders (other than IRD) impacting sensory/motor/cognitive function and eye diseases different from those of the study.
- Contraindications to TMS/EEG (e.g., metal implants, epilepsy).
- Non-correctable hearing or olfactory/gustatory deficits independent of IRD.

Inclusion criteria for controls:

- Age, sex, and handedness matched to the patient group.
- No history of ocular disease, neurological or psychiatric disorder, or sensory/motor impairment.
- Ability to provide written, informed consent.

Exclusion criteria - controls:

- Same as patient exclusion criteria.

### 2.5 Study Procedures and Assessments

Patients will complete a battery of five assessments. The duration of each assessment session is expected to vary by assessment type: approximately 1 hour for the genetic and family history interview, motor evaluation, chemosensory evaluation, and psycho-personological evaluation; and approximately 5 hours for the neurophysiological evaluation, including breaks. Patients will complete all assessments over two consecutive days. Sighted control participants will undergo the same assessment battery over two to four separate sessions within one month of enrolment. For both groups, the total duration of study-related testing is expected to be approximately 9-10 hours.

**Table 1.**
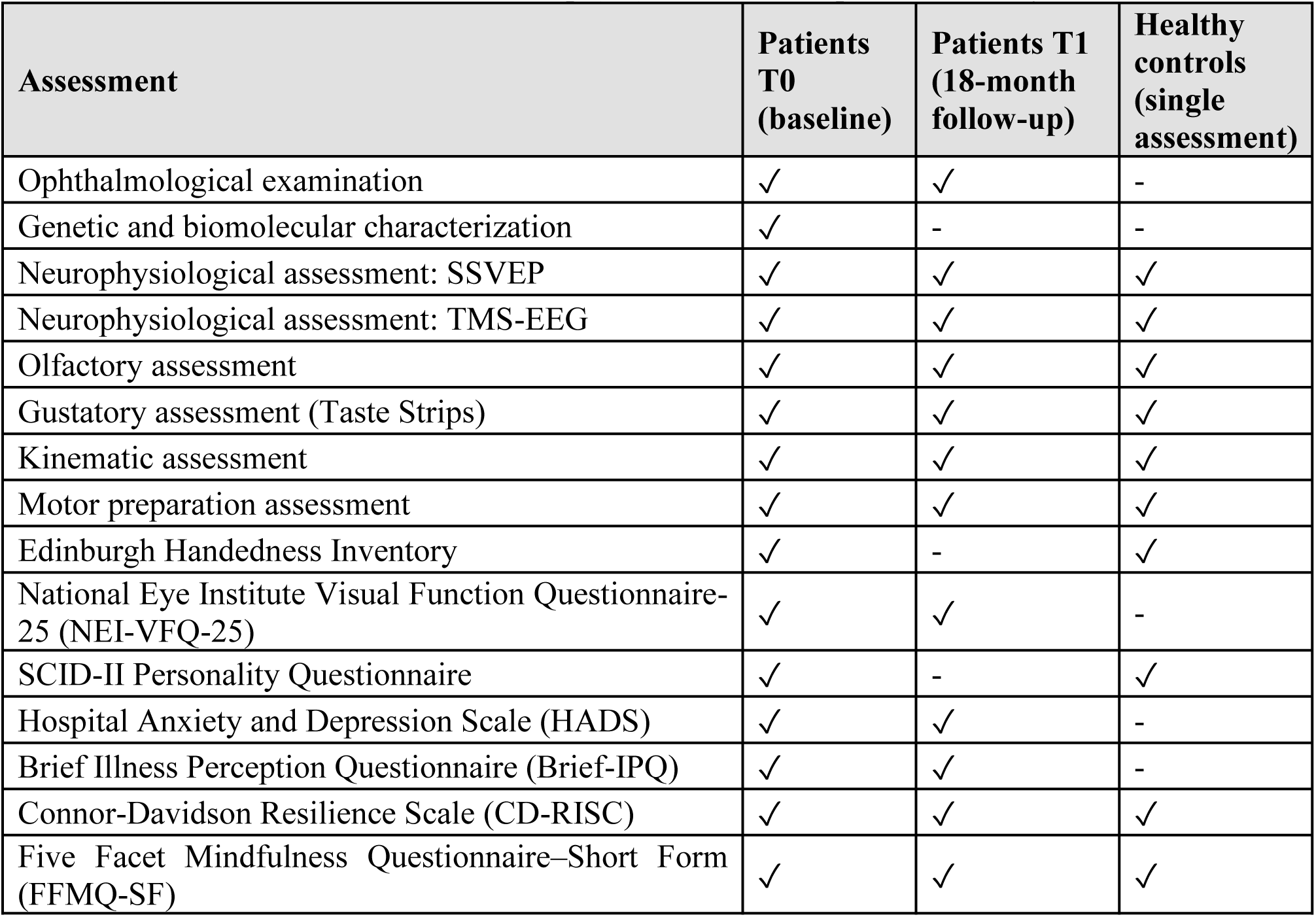
Assessment schedule across the five domains by timepoint and group. A check mark denotes an administered measure, and a hyphen denotes a measure not administered. Patients are assessed at baseline (T0) and at the 18-month follow-up (T1); controls complete the battery once.

| Assessment | Patients T0 (baseline) | Patients T1 (18-month follow-up) | Healthy controls (single assessment) |
| --- | --- | --- | --- |
| Ophthalmological examination | ✓ | ✓ | - |
| Genetic and biomolecular characterization | ✓ | - | - |
| Neurophysiological assessment: SSVEP | ✓ | ✓ | ✓ |
| Neurophysiological assessment: TMS-EEG | ✓ | ✓ | ✓ |
| Olfactory assessment | ✓ | ✓ | ✓ |
| Gustatory assessment (Taste Strips) | ✓ | ✓ | ✓ |
| Kinematic assessment | ✓ | ✓ | ✓ |
| Motor preparation assessment | ✓ | ✓ | ✓ |
| Edinburgh Handedness Inventory | ✓ | - | ✓ |
| National Eye Institute Visual Function Questionnaire-25 (NEI-VFQ-25) | ✓ | ✓ | - |
| SCID-II Personality Questionnaire | ✓ | - | ✓ |
| Hospital Anxiety and Depression Scale (HADS) | ✓ | ✓ | - |
| Brief Illness Perception Questionnaire (Brief-IPQ) | ✓ | ✓ | - |
| Connor-Davidson Resilience Scale (CD-RISC) | ✓ | ✓ | ✓ |
| Five Facet Mindfulness Questionnaire–Short Form (FFMQ-SF) | ✓ | ✓ | ✓ |

### 2.6 Ophthalmological Assessment

The biochemistry/molecular genetics team, in collaboration with the ophthalmology team, will identify patients with genetically confirmed IRDs, provide initial information about the study, and, upon patient acceptance, transfer the relevant clinical documentation to the University of Verona research team. Patients’ documentation will include comprehensive ophthalmological and genetic data to characterize the status of retinal degeneration and its functional impact at the time of assessment.

Patients will undergo a comprehensive ophthalmological evaluation, with most data obtained from routine clinical assessments performed as part of standard care; when necessary, missing or outdated information will be supplemented by additional clinical evaluations.

Clinical history and genetic characterization: A structured clinical history will be collected, including general medical and ocular history, disease stage and severity, age at symptom onset, and disease progression, as determined by longitudinal follow-up data. Family pedigree information and inheritance pattern classification will also be recorded. Genetic diagnosis will include the affected gene, allelic mutations, and pathogenicity classification according to the American College of Medical Genetics and Genomics (ACMG) criteria.

Visual function assessment: Visual function data will include best-corrected visual acuity, assessed monocularly in both eyes, refraction (sphere, cylinder, and axis), and intraocular pressure measured by tonometry.

Structural ophthalmological examination: Structural examination will include anterior segment evaluation by slit-lamp biomicroscopy; assessment of lens status, including phakic/pseudophakic status and lens transparency; and dilated fundus examination. Fundus description will include optic disc appearance, retinal vasculature, macular status, and peripheral retinal changes, including pigmentary deposits and atrophy patterns.

Multimodal retinal imaging and visual field assessment: Multimodal retinal imaging will include color fundus photography to document retinal pigmentary changes and vascular attenuation, optical Coherence Tomography (OCT) to assess outer retinal layer integrity, foveal morphology, and the extent of photoreceptor atrophy, and fundus autofluorescence to evaluate retinal pigment epithelium integrity and delineate areas of atrophy or abnormal autofluorescence. Microperimetry will be used to assess retinal sensitivity and correlate it with structural findings. Visual field assessment will be performed using the Humphrey Visual Field Analyzer (HFA Carl Zeiss Meditec, Dublin, CA, USA) with the 30-2 Swedish Interactive Threshold Algorithm (SITA)-Fast program. The assessment will document central and peripheral visual field function within the central 30°. It will include the Visual Field Index (VFI), Mean Deviation (MD), Pattern Standard Deviation (PSD), and Glaucoma Hemifield Test -GHT-results. Perimetry exam is critical for characterizing the pattern and extent of visual field loss, which may differ across IRD subtypes.

Self-administered Questionnaires: In addition to the consent procedure, each participant will complete the Edinburgh Handedness Inventory (Oldfield, 1971) to assess manual dominance. Patients will additionally complete the Italian version of the National Eye Institute Visual Function Questionnaire-25 (NEI VFQ-25, Mangione et al., 2001), a widely validated patient-reported outcome measure designed to quantify the subjective impact of visual impairment on daily functioning and vision-related quality of life (Mangione et al., 1998a, 1998b). It assesses multiple domains, including general vision, near and distance activities, peripheral and color vision, mental health related to visual problems, social functioning, dependency, and role limitations, thereby providing a comprehensive profile of the patient’s perceived visual disability and self-assessed functional limitations. Patients will also complete an additional instrument assessing the presence of visual anomalies and hallucination-like experiences (e.g., the Charles Bonnet Syndrome screening questionnaire; Cantin et al., 2018). All materials will be provided in paper format. Participants will be provided with a pen and seated comfortably at a table in a well-lit room to complete the questionnaires independently. A researcher remains present throughout the assessment to aid and clarify any ambiguous or unclear items as needed. For patients with compromised reading ability due to visual impairment, an adapted administration protocol will be implemented. The researcher will read aloud the entire informed consent document and all questionnaire forms. If the patient agrees to participate, they will be asked to sign the consent form. Subsequently, the researcher will verbally administer all questionnaire items and record the patient’s responses directly on the forms, without interpreting or influencing their answers.

### 2.7 Genetic and Biomolecular Assessment

Following the initial identification of the patient’s genotype in collaboration with the ophthalmology team, the biochemical/molecular genetics team will reconstruct the segregation pattern of the mutation, with reference to any known cases of familial recurrence. Characterization will proceed along two complementary lines:

1. Prior to the patient interview, based on the genetic screening data, an in silico structural model of the variant will be generated. When an experimentally solved structure of the target protein is available in the Protein Data Bank (Berman et al., 2000), this will be used as the primary template; otherwise, a predicted structure will be retrieved from the AlphaFold Protein Structure Database (Jumper et al., 2021). Mutant models will be generated and refined using the BioLuminate suite (Schrödinger, LLC, New York, NY) and compared with the corresponding wild-type structure to predict the mutation’s possible effects on measurable molecular parameters (e.g., protein stability, changes in affinity with known molecular partners), or to infer such effects based on the molecular pathway of the encoded protein. This model will be used to predict the mutation’s possible effects on measurable molecular parameters. The variant will be cross-checked against ClinVar (https://www.ncbi.nlm.nih.gov/clinvar/) and RetNet (https://retnet.org/) to retrieve any existing molecular-level annotations. If the variant is novel, a putative pathogenetic mechanism will be proposed based on the in silico predictions.
2. During the patient encounter, the proband’s clinical history and segregation data will be reconstructed, collecting detailed information on disease onset, symptoms, and family history, while excluding confounding genetic factors (e.g., mosaicism, uncertain genetic background). Finally, for patients carrying mutations in the same gene, a comparative structural analysis will be performed to rank variants by predicted molecular severity (e.g., degree of destabilization or altered binding relative to wild-type), providing a basis for correlations with the multimodal/functional study. The specific quantitative index used for this ranking will be defined based on the parameters available for each gene/protein family.

### 2.8 Neurophysiological Assessment

The neurophysiological assessment consists of two consecutive sessions designed to probe complementary aspects of visual system function in individuals with inherited retinal dystrophies. A first steady-state visual evoked potentials (SSVEPs) session involved EEG recordings aimed at quantifying how much visual information reaches the visual areas and evaluating the functional integrity and responsiveness of residual visual pathways under controlled visual stimulation. The second session combined TMS and EEG to measure TMS-evoked potentials (TEPs) and thereby assess the reactivity and effective connectivity of the early visual cortices (i.e., V1 and V2) and the whole brain in the absence of external visual input, assessing how the entire system works.

The two sessions will always be performed in the same order as SSVEPs followed by TMS–EEG, and will be separated by an extended break. This order is chosen to prevent any transient TMS-induced modulation of cortical modulation, such as short-lived potentiation or inhibition, from influencing visually driven EEG responses. Therefore, conducting the EEG session first ensures that the assessment of visually evoked activity is not biased by prior neuromodulatory effects.

#### 2.8.1 SSVEPs

The SSVEP paradigm follows an established frequency-tagging methodology (Norcia et al., 2015). The visual stimuli will be generated using a custom MATLAB script specifically developed for this study. All images will be presented on a 65-inch touch monitor (Smart Media) positioned 57 cm from the participant and rendered on a uniform gray background to minimize luminance artifacts and ensure high-contrast presentation across participants with varying levels of visual impairment. The stimulus will be a log-polar checkerboard subdivided into three concentric rings, each divided into eight angular sectors of 45°, yielding 24 independently stimulable sectors. Sector centers fall on the four cardinal meridians (horizontal and vertical) and the four diagonal meridians. This configuration stimulates retinal regions along the foveal-peripheral gradient, assessed via EEG, to determine where and how visual information, presented across different eccentricities and orientations, remained capable of triggering an entrained neural response (Hagler et al., 2009; Zhang et al., 2019) as a function of visual awareness.

The geometry of the ring follows from a single constraint: each ring should activate approximately equal cortical territory. This requires annular borders to follow a geometric progression in shifted eccentricity coordinates, as prescribed by the Rovamo–Virsu cortical magnification model (Rovamo & Virsu, 1979). Importantly, this model captures the average spatial-frequency tuning of the visual hierarchy, including extrastriate contributions beyond V1, and is therefore more appropriate for scalp-level SSVEP recordings than the purely striate model of Horton & Hoyt (1991), in which foveal dominance is far more extreme. The inner border is placed at E_min 2° to minimize contamination from fixation instability, a consideration relevant to the IRD population tested here. The outer border is set at E_max = 30°, chosen to encompass the full testable extent of the Humphrey perimetry. With E₂ = 3° as the Rovamo–Virsu offset parameter, the geometric ratio r = [(E_max + E₂)/(E_min + E₂)]^(1/3) = (33/5)^(1/3) ≈ 1.874 yields intermediate borders at 6.37° and 14.56°. Ring 1 thus spans 2°–6.37° (macular and parafoveal region), Ring 2 spans 6.37°–14.56° (mid-peripheral), and Ring 3 spans 14.56°–30° (far peripheral).

Check size is calibrated to match the spatial frequency tuning of the pattern-reversal VEP at each eccentricity. Harter (1970) demonstrated that the check size that elicits maximal VEP amplitude varies systematically with eccentricity: larger checks are required at greater eccentricities to drive the cortical representation optimally. Check size therefore scales linearly with (E + E₂), implemented via a log-polar indexing scheme in which the radial bin index increases logarithmically with eccentricity. This construction keeps checks approximately square at all eccentricities and ensures that the spatial frequency of the checkerboard decreases as (E + E₂)^(−1) across the full stimulus range, consistent with E₂ = 3°. At the geometric midpoint of each ring (3.57°, 9.65°, and 20.92°), the resulting check sizes are 16.1, 45.2, and 93.8 arcmin, corresponding to spatial frequencies of 1.86, 0.66, and 0.32 cpd, respectively.

The paradigm will be implemented using E-Prime 3.0 (Psychology Software Tools, Pittsburgh, USA). One sector at a time is driven by pattern reversal at 6 Hz (black-and-white alternation; Michelson contrast approximately 97%; black: 0.23 cd/m²; white: 82.5 cd/m²; background luminance: 47.7 cd/m²), while the remaining 23 sectors display a static low-contrast checkerboard (43.0–51.1 cd/m²). A central fixation point (radius 0.1°) is present throughout. Each pattern lasts 83.3 ms, and the entire stimulation trial (120 frames) lasts 10s (FLICKERING), followed by a 2s (STILL) static inter-trial interval.

The experiment comprises two pre-stimulation baseline conditions and six experimental blocks. Baselines will be recorded under both eyes-open and eyes-closed conditions using the same temporal structure as the stimulation trials, but without actual flickering (static gray pattern -STILL condition-). Each baseline lasts 2 minutes and includes pseudo-triggers equivalent in number to the stimulation trials (1440 markers per condition).

During the experimental session, each of the 24 sectors will be stimulated twice per block in pseudo-random order, with the constraint that repetitions of the same sector could not occur consecutively. Each block includes 48 FLICKERING sessions (24 sectors × 2 repetitions; total ≈ 8 min per block), followed by a short rest period. In total, six blocks will be presented, yielding 288 trials (12 repetitions per sector). During the flickering phase, the participant will keep their eyes fixed on the central black dot, which will always be superimposed on the images (both in the STILL and FLICKERING conditions), without moving their gaze towards the black-and-white sector. The experimental subject will be instructed to keep their eyes open throughout the visual stimulation and to limit blinking.

Participants could take self-paced breaks between blocks (∼ 1–2 min each) to minimize fatigue. The entire recording session lasts approximately 45 minutes, including baseline and stimulation periods. Accounting for the EEG setup (∼30 min) and task instructions (∼5 min), the total SSVEP procedure requires approximately 90 minutes per participant.

#### 2.8.2 EEG Acquisition, Preprocessing, and SSVEPs Analysis

EEG is recorded continuously throughout the experiment using a BrainVision amplifier (Brain Products GmbH, Gilching, Germany) with a 64-channel cap arranged according to the extended 10-10 system. Electrode impedance is kept below 5 kΩ throughout the session. Signals are digitized at 1000 Hz, referenced online to the right mastoid, and grounded at AFz.

Offline EEG preprocessing and analysis will be performed using EEGLAB (Delorme & Makeig, 2004) and custom MATLAB scripts, following established best practices for SSVEP research (Sánchez-López et al., 2019).

Raw EEG signals will be band-pass filtered between 0.01 and 49 Hz to remove slow drifts and high-frequency noise unrelated to cortical activity, while preserving the frequency components relevant for SSVEP responses. Data will then be sampled at 1000 Hz to achieve adequate temporal resolution for frequency-domain analysis. All preprocessing steps will be applied consistently across baseline and stimulation conditions to ensure comparability.

Artifact correction will be carried out using independent component analysis (ICA). Components associated with ocular artifacts (eye blinks and eye movements) will be identified by their scalp topography, temporal profile, and spectral characteristics, and removed prior to signal reconstruction. This step is essential to minimize non-neural contamination of electrodes.

Data will be segmented according to event markers. Epochs extended from 0 to +2000 ms relative to stimulation onset, providing a 2000 ms stimulation window flanked by stimulus periods. Mean time-domain baseline correction on the single epoch will be applied at this stage.

Automated epoch rejection will be applied using multiple statistical criteria: joint probability (epoch threshold: 5 SD; channel threshold: 3 SD), kurtosis (epoch: 5 SD; channel: 3 SD), amplitude threshold (-80 and 80 mV per channel), and linear trend detection (slope >50 µV/s, R² >0.3). For epochs with ≤6 problematic channels, spherical spline interpolation will be applied; epochs with >6 problematic channels will be rejected entirely. Channels flagged in >30% of epochs will be interpolated globally. Subsequently, EEG data will be divided according to experimental conditions and transformed into the frequency domain using Fast Fourier Transform (FFT). Steady-state visual evoked responses will be quantified by extracting spectral amplitudes and phases at the stimulation frequency (12 Hz) and its harmonics (24 Hz, 36 Hz) during the FLICKERING condition, which serve as reliable markers of stimulus-locked visual cortical synchronization in SSVEP paradigms (Norcia et al., 2015). The pipeline analysis is created to maximize signal-to-noise ratio (SNR) normalization and spectral comparability.

Preprocessing will be conducted at the whole-scalp level. In contrast, statistical analysis of SSVEP responses will focus on an occipital electrode cluster encompassing the primary visual areas (O1, Oz, and O2), where SSVEP responses are expected to be maximal.

#### 2.8.3 TMS-EEG

Single-pulse TMS (spTMS) will be employed to probe cortical excitability and connectivity. Stimulation will be delivered using a Magstim Rapid² stimulator (Magstim Company, Whitland, UK) equipped with a 70-mm figure-of-eight coil. Two cortical target sites will be investigated: the visual cortex (V1/V2; over left occipital electrode O1 and right occipital electrode O2) and the left primary motor cortex (M1).

Before stimulation, each participant will undergo 3D head digitization using an NDI Vicra optical tracking system (Northern Digital Inc., Waterloo, Canada), which uses infrared light to measure 3D position, to reconstruct an estimated individual MRI using Softaxic neuronavigation software (EMS, Bologna, Italy). This virtual MRI model will be used to guide coil positioning, ensuring reproducible stimulation across blocks and sessions.

For occipital (V1 & V2) stimulation, the coil will be held tangentially to the scalp, with the handle pointing upward and the coil plane oriented perpendicular to the skull surface to reduce participant discomfort. For motor cortex (M1) stimulation, the coil handle will be oriented downward, at 45° from the midline, and tilted until a clear motor evoked potential (MEP) is elicited in the contralateral hand muscles.

The resting motor threshold (rMT) will be determined in the dominant hand and defined as the minimum intensity required to elicit a MEP in the first dorsal interosseous (FDI) muscle of ≥50 μV in at least 5 out of 10 consecutive trials. The stimulation intensity for cortical targets will be set to 110% of the individual rMT.

For each cortical site (V1, V2, and M1), participants will receive 80 single pulses, divided into four blocks of 20 pulses each. Pulses will be delivered at randomized inter-stimulus intervals determined by the intrinsic TMS capacitor recharge time (5-7s), to prevent rhythmic entrainment and reduce anticipatory effects.

During stimulation, participants will be seated comfortably in a dimly lit room. They will be instructed to remain still, relax their facial muscles, and fixate on a central gray cross displayed on a black background on a computer screen positioned 57 cm in front of them throughout the entire session. No additional task or response will be required, minimizing cognitive load and isolating spontaneous cortical reactivity to stimulation. Short self-paced breaks will be provided between blocks to maintain attention and comfort.

EEG data will be recorded continuously throughout TMS delivery, with real-time data quality monitoring to detect and address artifacts. Each recording block lasts approximately 2 minutes, with rest breaks between blocks to minimize fatigue. The full TMS-EEG session, including coil placement and montage at each of the three stimulation sites, plus breaks, requires approximately 40 minutes.

#### 2.8.4 EEG Acquisition, Preprocessing, and TEP Extraction

EEG data will be recorded concurrently with TMS using the same 64-channel electrode system described for the SSVEP paradigm. Signals will be sampled at 5000 Hz with online reference at the right mastoid and ground at AFz.

Offline EEG preprocessing and TEP analysis will be performed using EEGLAB (Delorme & Makeig, 2004), the TMS-EEG Signal Analyzer (TESA) toolbox (Rogasch et al., 2017), and custom MATLAB scripts, following established best practices for TMS-EEG research.

Bad channels will be removed and interpolated. EEG data will be divided into 2000 ms epochs, with 1000 ms before TMS stimuli and 1000 ms after. The TMS pulse artifact (−1 to +8 ms) will be removed by cubic interpolation within a 20-ms time window. Then, EEG epochs will be downsampled to 1000 Hz, and bad trials will be manually rejected. The Independent Component Analysis (ICA) will first be used to remove TMS-related artifacts, and the time window from -2ms to 8ms will be replaced with cubic fitting.

Data will then be high-pass filtered at 1 Hz, low-pass filtered at 70 Hz, and notch-filtered at 48–52 Hz. The second step of ICA will be used to remove residual artifacts such as eye blinks or muscle noise. To improve ICA decomposition, the data between −2 and 8 ms following the TMS pulse will be replaced with constant-amplitude values prior to ICA and subsequently re-interpolated. Finally, re-references to the common average will be applied, and data will be baseline-corrected over the interval from –100 to –2ms. For subsequent analyses, the filtered data will be downsampled to 500 Hz.

#### 2.8.5 TEP Analysis and Visualization

TMS-evoked potentials (TEPs) will be computed as grand-averaged cortical responses time-locked to TMS pulse onset, calculated separately for each stimulation site (V1& V2, M1). TEPs will be analyzed within the first 50 ms by averaging the activity of 4 EEG channels around the stimulation site. For left occipital, electrodes O1, Oz, PO3, PO7 will be considered, while for right occipital electrodes O2, Oz, PO4, PO8 and for motor cortex the electrodes C1, C3, CP1, CP3. For both patients and healthy controls, the mean TEP amplitude will be calculated over two separate time windows centered on the first positive and first negative peaks.

To quantify overall cortical activation strength, the Global Mean Field Power (GMFP) will be computed as the spatial standard deviation of voltage across all electrodes at each time point (Lehmann & Skrandies, 1980; Esser et al., 2006).

GMFP will provide a reference-independent measure of synchronized cortical activity, with peaks that identify time points of maximal global activation, typically corresponding to canonical TEP components.

#### 2.8.6 Time-Frequency Analysis

Time–frequency analysis will be performed to investigate both oscillatory power and phase dynamics, providing a more comprehensive characterization of the underlying neurophysiological mechanisms. TMS-induced power will be calculated through the event-related spectral perturbation (ERSP) index, which quantifies the total power in the time and frequency domains relative to the pre-stimulation window. A Morlet wavelet will be used, and the ERSP will be evaluated as the average between 20-400 ms in three frequency bands: alpha (8-12 Hz), beta (13-30Hz), and gamma (31-45 Hz). This will allow comparison between the two groups in terms of evoked power over the overall time period.

The inter-trial phase consistency (ITPC) will also be employed to quantify the consistency across trials of the phase for a given time and frequency band.

#### 2.8.7 Effective Connectivity and Graph Analysis

To characterize brain functional connectivity following the TMS stimulation, the weighted phase lag index (wPLI) will be used. This metric quantifies phase synchronization between pairs of EEG electrodes across trials while minimizing the influence of common sources, such as volume conduction. A seed-based connectivity analysis will be conducted using electrodes surrounding the stimulation site for each target area (i.e., O1, O2, and C3). The connectivity between the selected channels and all others will be assessed within an early time window (i.e., 20-200ms) to investigate how stimulation shifts the brain network. Brain reorganization will also be evaluated through graph-based analysis. The properties of brain integration and segregation will be quantified using the following metrics: strength, clustering coefficient, characteristic path length, global and local efficiency, and small-world propensity.

### 2.9 Chemosensory Assessment

Chemosensory assessment will be conducted in both patients and controls at T0 and T1 using the same procedure. All testing will be conducted in a well-ventilated room with participants seated comfortably. Environmental conditions will be standardized to minimize external interference.

#### 2.9.1 Olfactory evaluation

Olfaction will be assessed through the Sniffin’ Sticks Extended Test (SSET) (Burghart Company, Holm, Germany), a well-known, validated test for comprehensive olfactory assessment in clinical and research contexts. It provides measurements of threshold (T), discrimination (D), and identification (I) in the olfactory domain, with normative data obtained from more than 9.000 subjects (Kobal et al., 1996; Hummel et al., 2007; Oleszkiewicz et al., 2019). Furthermore, immediately after the administration of the I test (consisting of 16 odorous items to identify, blue version), patients and controls will undergo an additional I test version with another 16 different odors (purple version) (Sorokowska et al., 2015). For both tests, participants will be asked to identify each odor before and after choosing among four possible verbal options, to make the task more cognitively demanding.

In particular, SSET consists of pen-like odor-dispensing devices, and the sum of the subtest scores (T, D, I) yields a global score (TDI) that defines a subject’s olfactory performance status. Normosmia (normal olfactory function) is established with a TDI score ≥ 30.5, hyposmia (reduced olfactory ability to detect odors) with a TDI score ≤30.75, and functional anosmia (total loss or minimal residual olfactory perception) with a TDI score ≤ 16.00. The T test shows the concentration at which one odor (butanone) is reliably detected and is performed using a single staircase procedure across 16 dilutions. The D test assesses the ability to distinguish odors by administering 16 triplets of odorous pens, each containing two identical odors and one different odor to find. The I test represents the ability to verbally identify an odor (overall 32 odors administered, without and right after, with verbal options to choose, as described previously). The global test is conducted with a forced-choice procedure.

#### 2.9.2 Gustatory evaluation

Gustation will be assessed by means of the “The Taste Strips Test” (TST) (Burghart Company, Holm, Germany), a validated test of gustatory function for the four basic taste qualities (sweet, sour, salty, bitter). This test consists of filter paper strips impregnated with different concentrations of the four basic tastes to identify. (Landis et al., 2009; Mueller et al., 2003).

In particular, the TST filter paper strips (8x2 cm) are impregnated with the following four concentrations for each taste quality: sweet (0.05, 0.1, 0.2, 0.4 g/mL sucrose), sour (0.05, 0.09, 0.165, 0.3 g/mL citric acid), salty (0.016, 0.04, 0.1, 0.25 g/mL NaCl), and bitter (0.0004, 0.0009, 0.0024, 0.006 g/mL quinine hydrochloride). Eighteen taste strips (four concentrations × four taste qualities + two blank strips) are presented in pseudorandomized order. Participants have to put the strip on the tongue for a whole mouth assessment and identify the taste quality (forced choice manner). Between every strip administration, the mouth needs to be rinsed with water to wash out the previous gustatory stimulus. Each correct answer yields one point. The total score (TST score) ranges from 0 to 16 (excluding the two blank strips). Normogeusia (normal gustatory performance) is defined with a score ≥9, hypogeusia (reduced gustatory performance) with a score <9. Patients with hypogeusia may show ageusia for certain taste qualities. A complete ageusia can be considered if the highest concentrations of all taste qualities are not detected (Mueller et al., 2003; Landis et al., 2009). For both olfactory and gustatory testing, the experimenter provided verbal instructions to ensure that participants with severe visual impairment could complete the assessments. Participants with preserved reading ability received written information and supplementary visual aids when needed.

Both chemosensory tests for olfaction and gustation will be easily administered to different kinds of visually impaired participants.

#### 2.9.3 Statistical Analysis on TDI and TST Scores and Subscales

Descriptive statistics (means, standard deviations, medians, interquartile ranges) will be computed for all chemosensory measures (TDI scores, individual T/D/I subscores, TST, and taste quality-specific scores).

- To assess whether visual function impairment affects chemosensory functioning, between-group comparisons of chemosensory performance will be conducted using independent-samples t-tests or Mann-Whitney U tests (depending on data distribution) to compare patients with hereditary retinal dystrophies against age- and sex-matched healthy controls.
- To assess whether the affected gene produced differential effects on olfactory and gustatory function, patients will be stratified according to genetic profile, and between-genotype comparisons will be performed using independent-samples t-tests or one-way ANOVA with post-hoc pairwise comparisons (Bonferroni-corrected) to identify genotype-specific patterns of chemosensory dysfunction.
- To assess whether there is a relationship between visual quality characterization and chemosensory function, Pearson or Spearman correlation will examine associations between chemosensory performance and clinical visual parameters, including: (a) age of symptom onset, (b) disease duration (current age minus onset age), and (c) subjective visual functioning as assessed by the NEI-VFQ-25.

All statistical tests will use a two-tailed alpha level of 0.05 with 95% confidence intervals. For Pearson or Spearman analyses, the strength and direction of linear associations will be quantified using correlation coefficients (r), with statistical significance evaluated via p-values. Correlation coefficients will be interpreted according to conventional guidelines: |r| = 0.10–0.29 (small effect), |r| = 0.30–0.49 (medium effect), |r| ≥ 0.50 (large effect) (Cohen, 1988). Effect sizes for ANOVA will be reported using Cohen’s d for t-tests and η² (eta-squared).

### 2.10 Motor Assessment

Assessing the ability to plan and execute appropriate actions is crucial in patients with retinal dystrophy, as vision is strongly involved in providing, for instance, the necessary information to estimate object properties (Cesari & Newell, 1999) and to guide movements in space (DiCaro et al., 2025; Jetter et al., 2026). Notably, every single motion to be smooth and precise needs to be preplanned, with the appropriate body posture and the correct muscle synergies; these are the so-called Anticipatory Postural Adjustments (APAs) (Aruin & Latash, 1996). Here, the aim is to test whether, in individuals with visual impairment, anticipatory action-related cues may partially compensate for the loss of visual information (Montani et al., 2026).

The experimental paradigm for motor assessment consists of two tasks, performed in separate blocks: i) in the “Grasping Task”, participants will be instructed to reach, grasp, and displace several objects of different sizes; ii) in the “APA Task”, participants will be required to self-perturb their quiet body posture by performing fast and explosive arm displacement.

Each participant will complete two blocks for each task, for a total of four blocks, with 28 trials per block.

#### 2.10.1 Scaling object size for grasping: Grasping Task

The subjects will be seated at a table. Their head position will be kept fixed with an adjustable chin rest, ensuring the participant’s eyes are aligned with the center of the stimulus, represented by a sphere on a pedestal located 30 cm from the participant.

The task for the participants will be to grasp, lift, and displace the object from one position to another. The spheres presented will be made in two sizes (Size factor). In half of the trials, they will be required to hold the pedestal with their left hand while performing the task, whereas in the other half, they will be required to keep their left hand on the table (Proprioceptive factor).

More specifically, the stimuli consist of two 3D-printed spheres (25 mm and 50 mm in diameter), placed at eye level on a pedestal.

The statistical design considered a between-subjects 2 × 2 × 2 factorial design, with Size (diameter 25 mm vs. 50 mm), Proprioceptive (hold the pedestal Yes vs. No), and Group (Patients vs. Healthy controls) as main factors.

The order of trials presenting the two stimulus sizes will be randomized within each experimental block, whereas the availability of proprioceptive cues will be manipulated across two separate blocks. The order of blocks will be counterbalanced across participants.

Kinematic data will be recorded using an electromagnetic tracker system (Viper Motion Tracking System, Polhemus, USA). The system will use a source to generate an electromagnetic field, which sensors will detect to compute the source’s position and orientation in real time. Hand movements will be tracked using three Micro Sensors 1.8: two placed on the participant’s right thumb and index finger, and one on the wrist. The 3D position of the hand will be captured at a sampling rate of 990 Hz.

For each trial, the kinematic recordings will be used to compute the following movement indices:

- Movement Time (MT): time elapsed from movement onset to object contact.
- Peak Velocity (PV): maximum wrist velocity during the reaching phase.
- Time to Peak Velocity (%MT): percentage of movement time elapsed before reaching peak velocity, providing information on the temporal organization of movement planning and online control.
- Peak Acceleration (PA): maximum acceleration reached during the transport phase.
- Peak Jerk (PJ): maximum (or normalized) jerk, used as an indicator of movement smoothness.
- Maximum Grip Aperture (MGA): maximum distance between thumb and index finger during reaching.
- Time to Maximum Grip Aperture (%MT): temporal occurrence of the maximum grip aperture relative to movement duration.
- Grip Aperture Scaling: modulation of maximum grip aperture as a function of object size.
- Perceptual Size Estimation Error: the difference between the perceived and the actual object size

#### 2.10.2 Anticipatory Postural Adjustments: APA task

Several studies have shown that Anticipatory Postural Adjustments (APAs) are altered in numerous neurological and musculoskeletal disorders, including Parkinson’s disease, stroke, cerebellar disorders, and other conditions affecting sensorimotor control (Cesari et al., 2022; Pascucci et al., 2023). Since visual information contributes to the construction and updating of internal models, visual impairment may also influence the generation and timing of APAs to maintain body balance.

Here, the self-perturbation paradigm (Aruin et al., 1996) will be used to investigate whether individuals with visual impairment exhibit altered anticipatory postural strategies and to determine the extent to which visual information contributes to feedforward postural control during voluntary movements.

The self-perturbation task employs a between-subjects 2 × 2 × 2 factorial design, with Arm Movement (Forward vs. Lateral), Eyes (Open vs. Closed), and Group (Patients vs. Healthy controls) as main factors. Trials involving the Arm Movements will be presented in a randomized order within each block, whereas the Eyes condition will be manipulated in two separate blocks. The order of blocks will be counterbalanced across participants.

At the beginning of each trial, participants will be instructed to maintain a relaxed posture, with their eyes either open or closed, and to await the experimenter’s signal specifying the required movement. Following this cue, participants will be asked to execute the specified movement as quickly as possible.

Muscle activity will be recorded using the Trigno Wireless Biofeedback System (Delsys, USA) with signals acquired from Trigno Avanti sensors. EMG sensors will be placed on five body muscles: tibialis anterior, soleus, rectus abdominis, erector spinae, and the back of the right hand to record movement onset and speed.

For each muscle, anticipatory postural adjustments (APAs) will be quantified by computing:

- EMG onset latency: onset of muscle activation detected using an automatic statistical algorithm based on a Moving Average Whitening Filter followed by an Approximate Generalized Likelihood Ratio (AGLR) detector.
- APA latency: temporal interval between EMG onset and kinematic movement onset.
- Movement onset: identified from the hand accelerometer signal.

### 2.11 Psychological and Personological Assessment

Assessing personality traits and psychological functioning in patients with retinal dystrophy is important, as vision impairment has been associated with poorer mental health (Lundeen et al., 2022). Conversely, protective psychological factors, such as resilience and mindfulness, may support more adaptive responses to chronic illness and disability (Cal et al., 2015; Greeson & Chin, 2019). Identifying both psychological challenges and personal strengths may help clinicians and researchers develop comprehensive clinical profiles that integrate physical and psychological functioning. Such profiles may contribute to a better understanding of patients’ adjustment to the condition. They may, in the longer term, help inform tailored support to promote well-being and quality of life and reduce the psychological burden associated with the disease.

#### 2.11.1 Personality Traits and Psychological Assessment Procedures

All assessments will be administered online via the LimeSurvey platform (version 6.10.3; LimeSurvey GmBH, Hamburg, Germany) after verifying that participants have sufficient reading autonomy to complete the questionnaires independently. Participants will complete the questionnaires on a computer, with an experimenter present and available to assist if needed (e.g., for clarification of item content or technical adjustments).

For patients whose visual impairment prevents independent reading, even with appropriate accommodations, the full test battery will be administered orally in a standardized interview format by a trained psychologist. This procedure will ensure accessibility while maintaining consistency and data quality across participants with different levels of visual impairment.

The following psychological domains will be assessed using validated Italian-language versions of standardized instruments:

- **SCID-II Personality Questionnaire** [First et al., 1997; First et al., 2003]. The SCID-II Personality Questionnaire is a 119-item self-report screening measure with dichotomous yes/no responses. It assesses the endorsement of personality-related features across several DSM-IV personality patterns, including avoidant, dependent, obsessive-compulsive, passive-aggressive, depressive, paranoid, schizotypal, schizoid, histrionic, narcissistic, and borderline patterns. The questionnaire will be used for descriptive personality profiling and not to establish clinical diagnoses.
- **Hospital Anxiety and Depression Scale (HADS)** [Zigmond & Snaith, 1983; Annunziata et al., 2011]. The HADS is a 14-item self-report instrument designed to assess symptoms of anxiety and depression. Items are rated on a 4-point response scale (0 = no symptoms/minimal intensity to 3 = severe symptoms), and the anxiety and depression subscale scores each range from 0 to 21. Scores are commonly interpreted as follows: 0–7, non-case; 8–10, possible case; and ≥11, probable case.
- **Brief Illness Perception Questionnaire (Brief-IPQ)** [Broadbent et al., 2006; Pain et al., 2006]. The Brief-IPQ is an 8-item questionnaire assessing cognitive and emotional representations of illness. Items are rated on 10-point scales and evaluate perceived consequences, timeline, personal control, treatment control, identity, concern, emotional response, and illness comprehensibility. After appropriate reverse scoring, an overall score ranging from 0 to 80 may be calculated, with higher scores indicating a more threatening perception of the illness. For descriptive purposes, scores may also be interpreted using the thresholds reported by Kuiper et al. [2022] in individuals with recently acquired spinal cord injury: <42, low perceived threat; 42–49, moderate perceived threat; and ≥50, high perceived threat. Because these thresholds were established in a different clinical population, the resulting categories should be interpreted with caution.
- **Connor-Davidson Resilience Scale (CD-RISC)** [Connor & Davidson, 2003; Ghisi et al., 2016]. The CD-RISC is a 25-item self-report questionnaire assessing an individual’s perceived capacity to cope with and recover from adversity, stress, and challenging life events. Items are rated on a 5-point Likert scale ranging from 0, “not true at all,” to 4, “true nearly all the time.” Total scores range from 0 to 100, with higher scores indicating greater resilience.
- **Five Facet Mindfulness Questionnaire–Short Form (FFMQ-SF)** [Baer et al., 2006; Bohlmeijer et al., 2011; Giovannini et al., 2014]. The FFMQ-SF is a 24-item self-report questionnaire providing a multidimensional assessment of mindfulness across five facets: observing, describing, acting with awareness, non-judging of inner experience, and non-reactivity to inner experience. Items are rated on a 5-point Likert scale ranging from 1, “never or very rarely true,” to 5, “very often or always true.” Facet scores and, where appropriate, a total score ranging from 24 to 120 will be reported, with higher scores indicating higher levels of mindfulness.

#### 2.11.2 Assessment Schedule

Psychological and personality assessments will be administered at T0 to both patients and healthy controls. A subset of the psychological measures will be repeated in the patient group at T1 to assess longitudinal changes in psychological functioning and adjustment.

- T0 assessments: Patient group: SCID-II, HADS, Brief-IPQ, CD-RISC, and FFMQ-SF. Control group: SCID-II, CD-RISC, and FFMQ-SF.
- T1 assessments: Patient group: HADS, Brief-IPQ, CD-RISC, and FFMQ-SF. Control group: Not assessed at T1

#### 2.11.3 Statistical Analysis of Questionnaire Scores

Descriptive statistics will be reported for all psychological and personality measures. Continuous scores will be summarized using means and standard deviations or medians and interquartile ranges, depending on their distribution, while categorical outcomes will be described using frequencies and percentages. At T0, measures administered to both patients and controls will be described separately for each group. Between-group differences will be explored using appropriate parametric or non-parametric tests, depending on the characteristics of the data. For measures repeated in patients at T1, changes over time will be examined using paired parametric or non-parametric tests, as appropriate. Given the anticipated small sample size, particular attention will be given to the magnitude and direction of observed differences, individual variability, and longitudinal change. Effect sizes and 95% confidence intervals will be reported where appropriate, and any inferential findings from the psychological analyses will be interpreted cautiously.

## 3. Discussion

This study protocol proposes a longitudinal multimodal approach to characterize functional changes associated with progressive visual loss in individuals with inherited retinal dystrophies. By combining ophthalmological and genetic characterization with neurophysiological, chemosensory, sensorimotor, and psychological assessments, the study is designed to characterize functional profiles and their variability across individuals.

The study allows both cross-sectional and longitudinal characterization. Baseline comparisons will characterize differences between individuals with IRDs and sighted controls across the assessed functional domains. In contrast, the 18-month follow-up will provide information on changes over the course of the disease within the same individuals. These longitudinal observations will be interpreted in relation to the clinical characteristics of visual impairment and, where sample size permits, to genotype-related differences.

A key feature of the protocol is that measures from different functional domains are collected in the same participants. Neurophysiological assessment will provide information on residual visual cortical responsiveness and cortical reactivity, while chemosensory and sensorimotor measures will characterize non-visual sensory and behavioral function. Psychological and personality-related measures will complement these assessments by describing individual differences in adjustment to the condition. This combined approach may therefore provide a more comprehensive characterization of the functional consequences of progressive visual impairment than approaches focusing on a single modality.

The genetic characterization included in the protocol is also relevant to interpreting inter-individual variability. Because IRDs are genetically heterogeneous and some disease-related genes may influence functions beyond the retina, differences observed in non-visual domains cannot necessarily be attributed solely to adaptation to visual loss. Examining functional measures in relation to genetic background and clinical characteristics may therefore help distinguish patterns primarily associated with visual impairment from those potentially influenced by genotype.

Several limitations should nevertheless be considered. IRDs are rare and genetically heterogeneous disorders, and the anticipated sample size may limit statistical power and the possibility of performing genotype-based subgroup analyses. For this reason, the study is explicitly exploratory, and particular attention will be given to effect sizes, individual variability, and the direction of longitudinal changes. In addition, the multimodal assessment battery is time- and attention-demanding and may affect the completeness of data collection, particularly among participants with more severe visual impairment. Finally, the 18-month follow-up represents only a limited interval within the generally slow course of retinal degeneration. It will therefore provide an initial characterization of longitudinal change rather than a complete description of long-term trajectories.

The protocol brings together clinical, genetic, neural, sensory, sensorimotor, and psychological information within the same individuals. This may contribute to a more individualized characterization of IRD-related functional profiles and provide a basis for future studies investigating monitoring and rehabilitation in progressive visual loss.

## 4. Ethics and Dissemination

The study protocol has been approved by the Ethics Committee of the University of Verona (CARP – Comitato di Approvazione della Ricerca sulla Persona; 08.R1/2024). All participants will provide written informed consent prior to participation, following a comprehensive explanation of the study procedures and the opportunity to ask questions. The study will be conducted in accordance with the Declaration of Helsinki, the General Data Protection Regulation (GDPR), and Italian Legislative Decree 196/2003 on personal data protection.

Findings will be disseminated through peer-reviewed publications and presentations at national and international scientific conferences. Anonymized, individual-level data and the analysis code will be deposited in a public repository or made available on reasonable request, subject to the data-protection provisions approved by the Ethics Committee. Given the rare-disease population included in the study, particular attention will be paid to minimizing the risk of participant re-identification when sharing individual-level data. Accessible summaries of the study findings will also be provided to participating patients and patient associations for inherited retinal dystrophies.

## Data Availability

All data produced in the present study will be made available online

## References

Annunziata, M. A., Muzzatti, B., & Altoè, G. (2011). Defining Hospital Anxiety and Depression Scale (HADS) structure by confirmatory factor analysis: a contribution to validation for oncological settings. Annals of Oncology, 22(10), 2330–2333. 10.1093/annonc/mdq750

Aruin, A. S., & Latash, M. L. (1996). Anticipatory postural adjustments during self-initiated perturbations of different magnitude triggered by a standard motor action. Electroencephalography and Clinical Neurophysiology, 101(6), 497–503. 10.1016/s0013-4694(96)95219-4

Avesani, A., Dal Cortivo, G., Asteriti, S., Targa, G., Veschetti, L., Marino, V., Biasi, A., Longo, C., Cisterna, B., Saran, K., Malerba, G., Foik, A. T., Cambiaghi, M., Cangiano, L., & Dell’Orco, D. (2026). Retinal network dysfunction precedes structural degeneration in severe GUCA1A cone-rod dystrophy. bioRxiv 2026.06.23.734025. 10.64898/2026.06.23.734025

Baer, R. A., Smith, G. T., Hopkins, J., Krietemeyer, J., & Toney, L. (2006). Using self-report assessment methods to explore facets of mindfulness. Assessment, 13(1), 27–45. 10.1177/1073191105283504

Berman, H. M., Westbrook, J., Feng, Z., Gilliland, G., Bhat, T. N., Weissig, H., Shindyalov, I. N., & Bourne, P. E. (2000). The Protein Data Bank. Nucleic Acids Research, 28(1), 235–242. 10.1093/nar/28.1.235

Bock, A. S., & Fine, I. (2014). Anatomical and functional plasticity in early blind individuals and the mixture of experts architecture. Frontiers in Human Neuroscience, 8, 971. 10.3389/fnhum.2014.00971

Bohlmeijer, E., ten Klooster, P. M., Fledderus, M., Veehof, M., & Baer, R. (2011). Psychometric properties of the Five Facet Mindfulness Questionnaire in depressed adults and development of a short form. Assessment, 18(3), 308–320. 10.1177/1073191111408231

Bonnet, C., & El-Amraoui, A. (2012). Usher syndrome (sensorineural deafness and retinitis pigmentosa): pathogenesis, molecular diagnosis and therapeutic approaches. Current Opinion in Neurology, 25(1), 42–49. 10.1097/WCO.0b013e32834ef8b2

Broadbent, E., Petrie, K. J., Main, J., & Weinman, J. (2006). The brief illness perception questionnaire. Journal of Psychosomatic Research, 60(6), 631–637. 10.1016/j.jpsychores.2005.10.020

Cantin, S., Duquette, J., Dutrisac, F., Ponton, L., Courchesne, M., de Abreu Cybis, W., Montisci, K., Wittich, W., & Wanet-Defalque, M.-C. (2018). Charles Bonnet syndrome: development and validation of a screening and multidimensional descriptive questionnaire. Canadian Journal of Ophthalmology, 54(3), 323–327. 10.1016/j.jcjo.2018.05.008

Castaldi, E., Cicchini, G. M., Falsini, B., Binda, P., & Morrone, M. C. (2019). Residual visual responses in patients with retinitis pigmentosa revealed by functional magnetic resonance imaging. Translational Vision Science & Technology, 8(6), 44. 10.1167/tvst.8.6.44

Castaldi E, Lunghi C, Morrone MC (2020). Neuroplasticity in adult human visual cortex. Neuroscience & Biobehavioral Reviews, 112: 542–552.

Cal, S. F., Sá, L. R. de, Glustak, M. E., & Santiago, M. B. (2015). Resilience in chronic diseases: A systematic review. Cogent Psychology, 2(1), 1024928. 10.1080/23311908.2015.1024928

Cesari, P., & Newell, K. M. (1999). The scaling of human grip configurations. Journal of Experimental Psychology: Human Perception and Performance, 25(4), 927–935. 10.1037/0096-1523.25.4.927

Cesari, P., Piscitelli, F., Pascucci, F., & Bertucco, M. (2022). Postural threat influences the coupling between anticipatory and compensatory postural adjustments in response to an external perturbation. Neuroscience, 490, 25–35. 10.1016/j.neuroscience.2022.03.005

Charbel Issa, P., Reuter, P., Kühlewein, L., Birtel, J., Gliem, M., Tropitzsch, A., Whitcroft, K. L., Bolz, H. J., Ishihara, K., MacLaren, R. E., Downes, S. M., Oishi, A., Zrenner, E., Kohl, S., & Hummel, T. (2018). Olfactory dysfunction in patients with CNGB1-associated retinitis pigmentosa. JAMA Ophthalmology, 136(7), 761–769. 10.1001/jamaophthalmol.2018.1621

Cohen, J. (1988). Statistical power analysis for the behavioral sciences (2nd ed.). Lawrence Erlbaum Associates.

Colombo, L., Maltese, P. E., Castori, M., El Shamieh, S., Zeitz, C., Audo, I., Zulian, A., Marinelli, C., Benedetti, S., Costantini, A., Bressan, S., Percio, M., Ferri, P., Abeshi, A., Bertelli, M., & Rossetti, L. (2021). Molecular epidemiology in 591 Italian probands with nonsyndromic retinitis pigmentosa and Usher syndrome. Investigative Ophthalmology & Visual Science, 62(2), 13. 10.1167/iovs.62.2.13

Connor, K. M., & Davidson, J. R. T. (2003). Development of a new resilience scale: The Connor-Davidson Resilience Scale (CD-RISC). Depression and Anxiety, 18(2), 76–82. 10.1002/da.10113

Delorme, A., & Makeig, S. (2004). EEGLAB: An open source toolbox for analysis of single-trial EEG dynamics including independent component analysis. Journal of Neuroscience Methods, 134(1), 9–21. 10.1016/j.jneumeth.2003.10.009

Dell’Orco, D., Sulmann, S., Zägel, P., Marino, V., & Koch, K. W. (2014). Impact of cone dystrophy-related mutations in GCAP1 on a kinetic model of phototransduction. Cellular and Molecular Life Sciences, 71(19), 3829–3840. 10.1007/s00018-014-1593-4

Di Caro, V., Cesari, P., Sala, F., & Cattaneo, L. (2025). The neural bases of the reach-grasp movement in humans: Quantitative evidence from brain lesions. Proceedings of the National Academy of Sciences, 122(10), e2419801122. 10.1073/pnas.2419801122

Duncan, J. L., Pierce, E. A., Laster, A. M., Daiger, S. P., Birch, D. G., Ash, J. D., Iannaccone, A., Flannery, J. G., Sahel, J. A., Zack, D. J., Zarbin, M. A., & the Foundation Fighting Blindness Scientific Advisory Board. (2018). Inherited retinal degenerations: current landscape and knowledge gaps. Translational Vision Science & Technology, 7(4), 6. 10.1167/tvst.7.4.6

Esser, S. K., Huber, R., Massimini, M., Peterson, M. J., Ferrarelli, F., & Tononi, G. (2006). A direct demonstration of cortical LTP in humans: a combined TMS/EEG study. Brain Research Bulletin, 69(1), 86–94. 10.1016/j.brainresbull.2005.11.003

First, M. B., Gibbon, M., Spitzer, R. L., Williams, J. B. W., & Benjamin, L. S. (1997). Structured Clinical Interview for DSM-IV Axis II Personality Disorders (SCID-II). American Psychiatric Press.

First MB, Gibbon M, Spitzer RL, Williams JBW, Benjamin LS. (2003). SCID-II: intervista clinica strutturata per i disturbi dell’Asse II del DSM-IV. Questionario di personalità. Mazzi F, Morosini P, De Girolamo G, Guaraldi GP, Italian adaptors. Firenze: Giunti Organizzazioni Speciali.

Geada, S., Teixeira-Marques, F., Teixeira, B., Carvalho, A. L., Lousan, N., Saraiva, J., Murta, J., Silva, R., Zanlonghi, X., Defoort-Dhellemmes, S., Smirnov, V., Dhaenens, C. M., Blanchet, C., Meunier, I., & Marques, J. P. (2023). Mutational spectrum, ocular and olfactory phenotypes of CNGB1-related RP-olfactory dysfunction syndrome in a multiethnic cohort. Genes, 14(4), 830. 10.3390/genes14040830

Ghisi, M., Bottesi, G., Re, A. M., Cerea, S., & Mammarella, I. C. (2016). Connor-Davidson Resilience Scale—Italian Version (CD-RISC) [Database record]. APA PsycTests. 10.1037/t61049-000

Gill, J. S., Georgiou, M., Kalitzeos, A., Moore, A. T., & Michaelides, M. (2019). Progressive cone and cone-rod dystrophies: clinical features, molecular genetics and prospects for therapy. British Journal of Ophthalmology, 103(5), 711–720. 10.1136/bjophthalmol-2018-313278

Giovannini, C., Giromini, L., Bonalume, L., Tagini, A., Lang, M., & Amadei, G. (2014). The Italian Five Facet Mindfulness Questionnaire: a contribution to its validity and reliability. Journal of Psychopathology and Behavioral Assessment, 36, 415–423. 10.1007/s10862-013-9403-0

Greeson, J. M., & Chin, G. R. (2019). Mindfulness and physical disease: A concise review. Current Opinion in Psychology, 28, 204–210. 10.1016/j.copsyc.2018.12.014

Hagler, D. J. Jr., Halgren, E., Martinez, A., Huang, M., Hillyard, S. A., & Dale, A. M. (2009). Source estimates for MEG/EEG visual evoked responses constrained by multiple, retinotopically-mapped stimulus locations. Human Brain Mapping, 30(4), 1290–1309. 10.1002/hbm.20597

Hamel, C. (2006). Retinitis pigmentosa. Orphanet Journal of Rare Diseases, 1, 40. 10.1186/1750-1172-1-40

Harter, M. R., & White, C. T. (1970). Evoked cortical responses to checkerboard patterns: Effect of check-size as a function of visual acuity. Electroencephalography and Clinical Neurophysiology, 28(1), 48–54. 10.1016/0013-4694(70)90007-6

Hartong, D. T., Berson, E. L., & Dryja, T. P. (2006). Retinitis pigmentosa. The Lancet, 368(9549), 1795–1809. 10.1016/S0140-6736(06)69740-7

Horton, J. C., & Hoyt, W. F. (1991). The representation of the visual field in human striate cortex. A revision of the classic Holmes map. Archives of Ophthalmology, 109(6), 816–824. PMID: 2043069.

Hummel, T., Kobal, G., Gudziol, H., & Mackay-Sim, A. (2007). Normative data for the “Sniffin’ Sticks” including tests of odor identification, odor discrimination, and olfactory thresholds: an upgrade based on a group of more than 3,000 subjects. European Archives of Oto-Rhino-Laryngology, 264(3), 237–243. 10.1007/s00405-006-0173-0

Jetter, C., He, C., Tu, A. S., Starrett, M. J., Chrastil, E. R., & Hegarty, M. (2026). Map-based navigation: Individual differences in perspective taking and path integration. Neuropsychologia, 229, 109476. 10.1016/j.neuropsychologia.2026.109476

Jumper, J., Evans, R., Pritzel, A., Green, T., Figurnov, M., Ronneberger, O., et al. (2021). Highly accurate protein structure prediction with AlphaFold. Nature, 596(7873), 583–589. 10.1038/s41586-021-03819-2

Karstensen, H. G., Mang, Y., Fark, T., Hummel, T., & Tommerup, N. (2015). The first mutation in CNGA2 in two brothers with anosmia. Clinical Genetics, 88(3), 293–296. 10.1111/cge.12491

Kobal, G., Hummel, T., Sekinger, B., Barz, S., Roscher, S., & Wolf, S. (1996). ”Sniffin’ Sticks”: a new test for olfactory performance. Rhinology, 34(4), 222–226. PMID: 9050101.

Kolarik, A., & Moore, B. C. J. (2024). Principles governing the effects of sensory loss on human abilities: An integrative review. Neuroscience & Biobehavioral Reviews, 169, 105986. 10.1016/j.neubiorev.2024.105986

Kuiper, H., van Leeuwen, C. M. C., Stolwijk-Swüste, J. M., & Post, M. W. M. (2022). Reliability and validity of the Brief Illness Perception Questionnaire (B-IPQ) in individuals with a recently acquired spinal cord injury. Clinical Rehabilitation, 36(4), 550–557. 10.1177/02692155211061813

Landis, B. N., Welge-Luessen, A., Brämerson, A., Bende, M., Mueller, C. A., Nordin, S., & Hummel, T. (2009). ”Taste Strips” – A rapid, lateralized, gustatory bedside identification test based on impregnated filter papers. Journal of Neurology, 256(2), 242–248. 10.1007/s00415-009-0088-y

Lehmann, D., & Skrandies, W. (1980). Reference-free identification of components of checkerboard-evoked multichannel potential fields. Electroencephalography and Clinical Neurophysiology, 48(6), 609–621. 10.1016/0013-4694(80)90419-8

Lundeen, E. A., Saydah, S., Ehrlich, J. R., & Saaddine, J. (2022). Self-reported vision impairment and psychological distress in U.S. adults. Ophthalmic Epidemiology, 29(2), 171–181. 10.1080/09286586.2021.1918177

Mangione, C. M., Lee, P. P., Pitts, J., Gutierrez, P., Berry, S., & Hays, R. D. (1998a). Psychometric properties of the National Eye Institute Visual Function Questionnaire (NEI-VFQ). Archives of Ophthalmology, 116(11), 1496–1504.

Mangione, C. M., Berry, S., Spritzer, K., Janz, N. K., Klein, R., Owsley, C., & Lee, P. P. (1998b). Identifying the content area for the 51-item National Eye Institute Visual Function Questionnaire: results from focus groups with visually impaired persons. Archives of Ophthalmology, 116(2), 227–233.

Mangione, C. M., Lee, P. P., Gutierrez, P. R., Spritzer, K., Berry, S., & Hays, R. D. (2001). Development of the 25-item National Eye Institute Visual Function Questionnaire. Archives of Ophthalmology, 119(7), 1050–1058. 10.1001/archopht.119.7.1050

Marino, V., Dal Cortivo, G., Oppici, E., Maltese, P. E., D’Esposito, F., Manara, E., Ziccardi, L., Falsini, B., Magli, A., Bertelli, M., & Dell’Orco, D. (2018). A novel p.(Glu111Val) missense mutation in GUCA1A associated with cone-rod dystrophy leads to impaired calcium sensing and perturbed second messenger homeostasis in photoreceptors. Human Molecular Genetics, 27(24), 4204–4217. 10.1093/hmg/ddy311

Mathur, P., & Yang, J. (2015). Usher syndrome: Hearing loss, retinal degeneration and associated abnormalities. Biochimica et Biophysica Acta (BBA) – Molecular Basis of Disease, 1852(3), 406–420. 10.1016/j.bbadis.2014.11.020

McEwen, D. P., Koenekoop, R. K., Khanna, H., Jenkins, P. M., Lopez, I., Swaroop, A., & Martens, J. R. (2007). Hypomorphic CEP290/NPHP6 mutations result in anosmia caused by the selective loss of G proteins in cilia of olfactory sensory neurons. Proceedings of the National Academy of Sciences, 104(39), 15917–15922. 10.1073/pnas.0704140104

Montani, V., Pascucci, F., Colombari, E., Savazzi, S., & Cesari, P. (2026). Visual awareness of stimulus features shapes motor control through action end-state comfort. Scientific Reports, 16(1), 10801. 10.1038/s41598-026-43752-w

Mueller, C., Kallert, S., Renner, B., Stiassny, K., Temmel, A. F., Hummel, T., & Kobal, G. (2003). Quantitative assessment of gustatory function in a clinical context using impregnated “taste strips”. Rhinology, 41(1), 2–6.

Norcia, A. M., Appelbaum, L. G., Ales, J. M., Cottereau, B. R., & Rossion, B. (2015). The steady-state visual evoked potential in vision research: A review. Journal of Vision, 15(6), 4. 10.1167/15.6.4

Oldfield, R. C. (1971). Edinburgh Handedness Inventory [Database record]. APA PsycTests. 10.1037/t23111-000

Oleszkiewicz, A., Schriever, V. A., Croy, I., Hähner, A., & Hummel, T. (2019). Updated Sniffin’ Sticks normative data based on an extended sample of 9139 subjects. European Archives of Oto-Rhino-Laryngology, 276(3), 719–728. 10.1007/s00405-018-5248-1

Pain D, Miglioretti M, Angelino E. (2006). Sviluppo della versione italiana del Brief-IPQ (Illness Perception Questionnaire, short version), strumento psicometrico per lo studio delle rappresentazioni di malattia. Psicol Salute. 1:81–89.

Pascucci, F., Cesari, P., Bertucco, M., & Latash, M. L. (2023). Postural adjustments to self-triggered perturbations under conditions of changes in body orientation. Experimental Brain Research, 241(8), 2163–2177. 10.1007/s00221-023-06671-0

Rogasch, N. C., Sullivan, C., Thomson, R. H., Rose, N. S., Bailey, N. W., Fitzgerald, P. B., Farzan, F., & Hernandez-Pavon, J. C. (2017). Analysing concurrent transcranial magnetic stimulation and electroencephalographic data: A review and introduction to the open-source TESA software. NeuroImage, 147, 934–951. 10.1016/j.neuroimage.2016.10.031

Rovamo, J., & Virsu, V. (1979). An estimation and application of the human cortical magnification factor. Experimental Brain Research, 37(3), 495–510. 10.1007/BF00236819

Sailani, M. R., Jingga, I., MirMazlomi, S. H., Bitarafan, F., Bernstein, J. A., & Snyder, M. P. (2017). Isolated congenital anosmia and CNGA2 mutation. Scientific Reports, 7(1), 2667. 10.1038/s41598-017-02947-y

Sanchez-Lopez J, Pedersini CA, Di Russo F, Cardobi N, Fonte C, Varalta V, Prior M, Smania N, Savazzi S, Marzi CA (2019). Visually evoked responses from the blind field of hemianopic patients. Neuropsychologia, 128: 127–139.

Sorokowska, A., Schriever, V. A., Gudziol, V., Hummel, C., Hähner, A., Iannilli, E., Sinding, C., Aziz, M., Seo, H. S., Negoias, S., & Hummel, T. (2015). Changes of olfactory abilities in relation to age: odor identification in more than 1400 people aged 4 to 80 years. European Archives of Oto-Rhino-Laryngology, 272(8), 1937–1944. 10.1007/s00405-014-3263-4

Wright, A. F., Chakarova, C. F., Abd El-Aziz, M. M., & Bhattacharya, S. S. (2010). Photoreceptor degeneration: genetic and mechanistic dissection of a complex trait. Nature Reviews Genetics, 11(4), 273–284. 10.1038/nrg2717

Zhang, N., Liu, Y., Yin, E., et al. (2019). Retinotopic and topographic analyses with gaze restriction for steady-state visual evoked potentials. Scientific Reports, 9, 4472. 10.1038/s41598-019-41158-5

Zigmond, A. S., & Snaith, R. P. (1983). The Hospital Anxiety and Depression Scale. Acta Psychiatrica Scandinavica, 67(6), 361–370. 10.1111/j.1600-0447.1983.tb09716.x

